# Avian Influenza Virus Infections in Felines: A Systematic Review of Two Decades of Literature

**DOI:** 10.1101/2024.04.30.24306585

**Authors:** Kristen K. Coleman, Ian G. Bemis

**Affiliations:** Department of Global, Environmental, and Occupational Health, University of Maryland School of Public Health, College Park, MD, 20742, USA; Department of Veterinary Medicine, College of Agriculture and Natural Resources, University of Maryland, College Park, MD, 20742, USA

**Keywords:** avian influenza virus, cross-species transmission, zoonosis, felines, cats, avian influenza pandemic, One Health

## Abstract

As an avian influenza virus (AIV) panzootic is underway, the threat of a human pandemic is emerging. Infections among mammalian species in frequent contact with humans should be closely monitored. One mammalian family, the Felidae, is of particular concern. Domestic cats are susceptible to AIV infection and provide a potential pathway for zoonotic spillover to humans. Here, we provide a systematic review of the scientific literature to describe the epidemiology and global distribution of AIV infections in felines reported from 2004 – 2024. We identified 607 AIV infections in felines, including 302 associated deaths, comprising 18 countries and 12 felid species. We observed a drastic flux in the number of AIV infections among domestic cats in 2023 and 2024, commensurate with the emergence of H5N1 clade 2.3.4.4b. We estimate that this phenomenon is underreported in the scientific literature and argue that increased surveillance among domestic cats is urgently needed.

## Introduction

Avian influenza virus (AIV) is an important and emerging zoonotic and reverse-zoonotic viral pathogen with pandemic potential. AIV has caused substantial disruptions to food supply chains, resulting in large economic losses in the poultry industry, as well as disruptions to regional and global food security. As avian influenza has a high mortality rate, an AIV pandemic could result in substantially more human illness and death than recent pandemics. While current surveillance is focused on AIV as an emerging pathogen in U.S. dairy cattle, infections among other susceptible domestic mammalian species have received comparatively little attention. In particular, AIV infections in free-roaming and farm-associated domestic cats are rising in the U.S. [1,2], but overall surveillance efforts among felines appear sparse.

Domestic cats are a popular human companion animal and thus provide a potential pathway for zoonotic spillover of avian influenza viruses to and from humans. Avian influenza in felines is often fatal, although subclinical infections have been reported [3,4]. Felines prey on wild birds and mammals and may serve as a host for AIV adaptation to mammals. Feline-to-feline transmission of AIV has been demonstrated experimentally [5,6], and real-world outbreaks among felines resulting in human infections have been reported [7–9]. To provide a comprehensive background for the assessment of the current risk, as well as to bring awareness to the recurring phenomenon of AIV infection in felids, we performed a systematic review of AIV infections in felines reported in the peer-reviewed scientific literature from 2004 through 2024. In this review, we sought to characterize the epidemiology and global distribution of AIV infections in felid species of domestic and wild origin reported over time, including information on AIV subtypes and clades identified.

## Methods

### Electronic literature search and article selection

We conducted systematic electronic searches for peer-reviewed articles in the English scientific literature to identify eligible studies. Our search strategy is presented in Supplementary Table 1. A study was eligible to be included if the following criteria were met: (1) avian influenza virus infection (including the abbreviation AIV) in “felines”, “felids”, or “cats” mentioned in the abstract, main text, or within keywords, including Pantherinae species names (i.e., lion, tiger, jaguar, leopard, snow leopard) and Felinae species names (e.g., cheetah, caracal, cat, wildcat, jaguarundi, ocelot, oncilla, tigrillo, kodkod, güiña, margay, southern tigrina, serval, lynx, bobcat, manul, cougar, puma, and catamount), and (2) surveillance/epidemiological studies, case reports, or laboratory/bioinformatics studies of animal samples (or data from animal samples) that report original/primary evidence of avian influenza virus detection in felines/cats within the abstract or main text. We excluded studies where: (1) sound detection methods for the identification of avian influenza virus were not used, (2) original/primary evidence of avian influenza virus detection was not reported (e.g., genomic studies of avian influenza virus specimens already reported in the peer-reviewed literature), (3) only experimental infection studies were performed, and (4) publication was after December 2024. Note that genomic studies of previously collected avian influenza virus specimens were included if the specimens were not reported elsewhere in the peer-reviewed scientific literature. Studies selected for inclusion are listed in Table 1.

**Table 1.** Publications included in our systematic review of avian influenza virus infections in felines published from 2004 – 2024, including country, feline species, influenza subtypes, clades, and year(s) of study.

| Reference | Published | Country | Feline species | Subtype(s) | Clade | Study year(s) |
| --- | --- | --- | --- | --- | --- | --- |
| Keawcharoen *et al.* [53] | 2004 | Thailand | Tiger (*Panthera tigris*) & leopard (*P. pardus*) | H5N1 |  | 2003 |
| Thanawongnuwech *et al.* [7] | 2005 | Thailand | Tiger (*P. tigris*) | H5N1 |  | 2004 |
| Editorial Team [56] | 2006 | Germany | Domestic cat (*Felis catus*) | H5N1 |  | 2006 |
| Songserm *et al.* [57] | 2006 | Thailand | Domestic cat (*F. catus*) | H5N1 |  | 2004 |
| Yingst *et al.* [58] | 2006 | Iraq | Domestic cat (*F. catus*) | H5N1 |  | 2006 |
| Leschnik *et al.* [3] | 2007 | Austria | Domestic cat (*F. catus*) | H5N1 |  | 2006 |
| Klopfleisch *et al.* [59] | 2007 | Germany | Domestic cat (*F. catus*) | H5N1 |  | 2006 |
| Mushtaq *et al.* [60] | 2008 | China | Tiger (*P. tigris*) | H5N1 | clade 2.2 | 2005 |
| Lam *et al.* [61] | 2008 | Indonesia | Domestic cat (*F. catus*) | H5N1 |  | 2005-2007 |
| Desvaux *et al.* [54] | 2009 | Cambodia | Tiger (*P. tigris*), Asian golden cat (*Catopuma temminckii*), leopard (*P. pardus*), clouded leopard (*Neofelis nebulosa*) | H5N1 | clade 1 | 2004 |
| El-Sayed *et al.* [62] | 2013 | Egypt | Domestic cat (*F. catus*) | H5N1 |  |  |
| He *et al.* [63] | 2015 | China | Bengal tiger (*P. tigris tigris*) | H5N1 |  | 2013 |
| Zhou *et al.* [64] | 2015 | China | Domestic cat (*F. catus*) | H5N1 and H9N2 |  | 2010-2012 |
| Sun *et al.* [65] | 2015 | China | Domestic cat (*F. catus*) | H5N1 |  | 2013 |
| Yu *et al.* [66] | 2015 | China | Domestic cat (*F. catus*) | H5N6 |  | 2014 |
| Zhao *et al.* [67] | 2015 | China | Domestic cat (*F. catus*) | H5N1 | clade 2.3.2 |  |
| Hu *et al.* [68] | 2016 | China | Tiger (*P. tigris*) | H5N1 | 2.3.4.4e, 2.3.2.1b, and 2.3.2.1c | 2014 and 2015 |
| Chen *et al.* [69] | 2016 | China | Lion (*P. leo*) | H5N1 | clade 2.3.2.1c | 2016 |
| Newbury *et al.* [70] | 2017 | USA | Domestic cat (*F. catus*) | H7N2 |  | 2016 |
| Cao *et al.* [71] | 2017 | China | Domestic cat (*F. catus*) | H5N6 | novel reassortants | 2016 |
| Lee *et al.* [8] | 2017 | USA | Domestic cat (*F. catus*) | H7N2 |  | 2016 |
| Mahardika *et al.* [27] | 2018 | Indonesia | Domestic cat (*F. catus*) | H5 |  | 2006 |
| Kim *et al.* [72] | 2018 | Korea | Leopard cat (*Prionailurus bengalensis*) | H5 |  | 2011-2016 |
| Wang *et al.* [73] | 2018 | China | Siberian tiger (*P. tigris altaica*) | H5 and H9 |  | 2015-2016 |
| Lee *et al.* [74] | 2018 | South Korea | Domestic cat (*F. catus*) | H5N6 |  | 2016 |
| Sangkachai *et al.* [75] | 2019 | Thailand | Bengal tiger (*P. tigris tigris*) | H5N1 |  | 2012 and 2013 |
| Soilemetzidou *et al.* [26] | 2020 | Namibia | Caracal (*Caracal caracal*) | H1, H3, H5, H7, H8, H9, H11, H13, H14, and H16 |  | 2009-2013 |
| Zhao *et al.* [76] | 2021 | China | Domestic cat (*F. catus*) |  |  | 2017 |
| Bao *et al.* [77] | 2022 | China | Domestic cat (*F. catus*) | H3N8 | reassortant | 2022 |
| Tammiranta *et al.* [28] | 2023 | Finland | Lynx (*Lynx lynx*) | H5N1 | clade 2.3.4.4b | 2022 |
| Briand *et al.* [29] | 2023 | France | Domestic cat (*F. catus*) | H5N1 | clade 2.3.4.4b | 2022 |
| Rabalski *et al.* [30] | 2023 | Poland | Domestic cat (*F. catus*) & caracal (*C. caracal*) | H5N1 | clade 2.3.4.4b | 2023 |
| Domańska-Blicharz *et al.* [31] | 2023 | Poland | Domestic cat (*F. catus*) | H5N1 | clade 2.3.4.4b | 2023 |
| Sillman *et al.* [1] | 2023 | USA | Domestic cat (*F. catus*) | H5N1 | clade 2.3.4.4b | 2023 |
| Moreno *et al.* [4] | 2023 | Italy | Domestic cat (*F. catus*) | H5N1 | clade 2.3.4.4b | 2023 |
| Szaluś-Jordanow *et al.* [32] | 2023 | Poland | Domestic cat (*F. catus*) | H5N1 |  | 2023 |
| Yang *et al.* [78] | 2023 | China | Domestic cat (*F. catus*) | H9N2 | clade C1.2 | 2018 |
| Elsmo *et al.* [33] | 2023 | USA | Bobcat (*Lynx rufus*) | H5N1 | clade 2.3.4.4b | 2022 |
| Cruz *et al.* [34] | 2023 | Peru | Lion (*P. leo*) | H5N1 | clade 2.3.4.4b | 2023 |
| Villanueva-Saz *et al.* [40] | 2024 | Spain | Domestic cat (*F. catus*) | H5 |  | 2022-2023 |
| Lee *et al.* [35] | 2024 | South Korea | Domestic cat (*F. catus*) | H5N1 | clade 2.3.4.4b | 2023 |
| Kang *et al.* [36] | 2024 | South Korea | Domestic cat (*F. catus*) | H5N1 | clade 2.3.4.4b | 2023 |
| Burrough *et al.* [2] | 2024 | USA | Domestic cat (*F. catus*) | H5N1 | clade 2.3.4.4b | 2024 |
| Cavicchio *et al.* [79] | 2024 | Italy | Domestic cat (*F. catus*) | H5 |  | 2021-2022 |
| Duijvestijn *et al.* [41] | 2024 | Netherlands | Domestic cat (*F. catus*) | H5N1 | clade 2.3.4.4b | 2020-2023 |
| Caserta *et al.* [37] | 2024 | USA | Domestic cat (*F. catus*) | H5N1 | clade 2.3.4.4b | 2024 |
| Chothe *et al.* [38] | 2024 | USA | Domestic cat (*F. catus*) | H5N1 | clade 2.3.4.4b | 2024 |
| Mainenti *et al.* [80] | 2024 | USA | Domestic cat (*F. catus*) | H5N1 | clade 2.3.4.4b | 2024 |

### Data extraction and statistical analysis

Data such as number of avian influenza virus infections, subtype, clade, location, intra- or cross-species transmission, disease characteristics, outcomes, and species infected were extracted from full-text articles using a data collection form. Data were imported to STATA version 15.1 (StataCorp, College Station, TX, USA) for descriptive statistical analyses.

## Results

Our electronic search generated 207 articles through PubMed and 207 through Scopus. We identified a total of 18 experimental feline infection studies [5,6,10–25] and excluded them from our review. After further screening, we included a total of 48 articles in our review (Table 1), two of which were identified manually through article references [26,27]. Of these articles, 33 used reverse transcription polymerase chain reaction (RT-PCR) alone or as a preparation for sequencing, and 13 used serology as their primary method of detection. See Figure 1 for a PRISMA consort diagram of article screening and selection.

**Figure 1.**
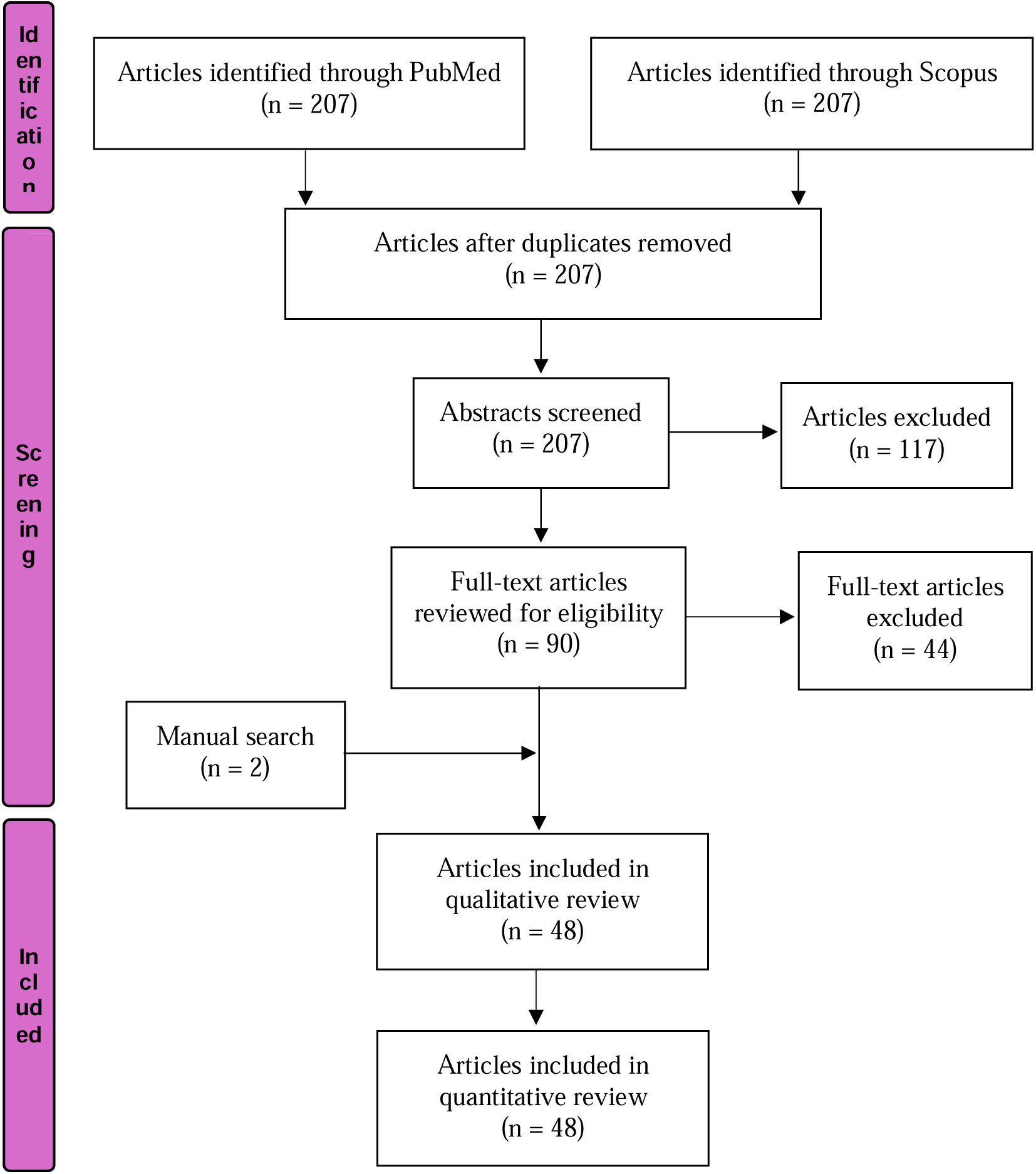
PRISMA flow diagram for identification, screening and review of the peer-reviewed scientific literature on avian influenza virus infections in felines published from 2004 – 2024

### Avian influenza virus infections in felines reported over time

From 2004 – 2024, approximately 607 avian influenza virus infections in felines, including 302 associated feline deaths, were reported in the English scientific literature. In total, 184 of these infections were identified by serological methods alone, while the rest were identified by RT-PCR or viral culture followed by genome sequencing. A total of 62.6% (380/607) of the reported feline cases were domestic cats. The annual number of articles reporting avian influenza virus infections in felines drastically increased in 2023 (Figure 2), with a recent spike in the total number of domestic cat infections and deaths reported in 2023 and 2024 (Figures 3 and 4). This spike is commensurate with the emergence and increased spread of avian influenza virus H5N1 clade 2.3.4.4b among birds and mammals. As publication year may not be representative of the actual year(s) in which reported feline infections occurred, we provide data on the specific year(s) samples were collected (Table 1).

**Figure 2.**
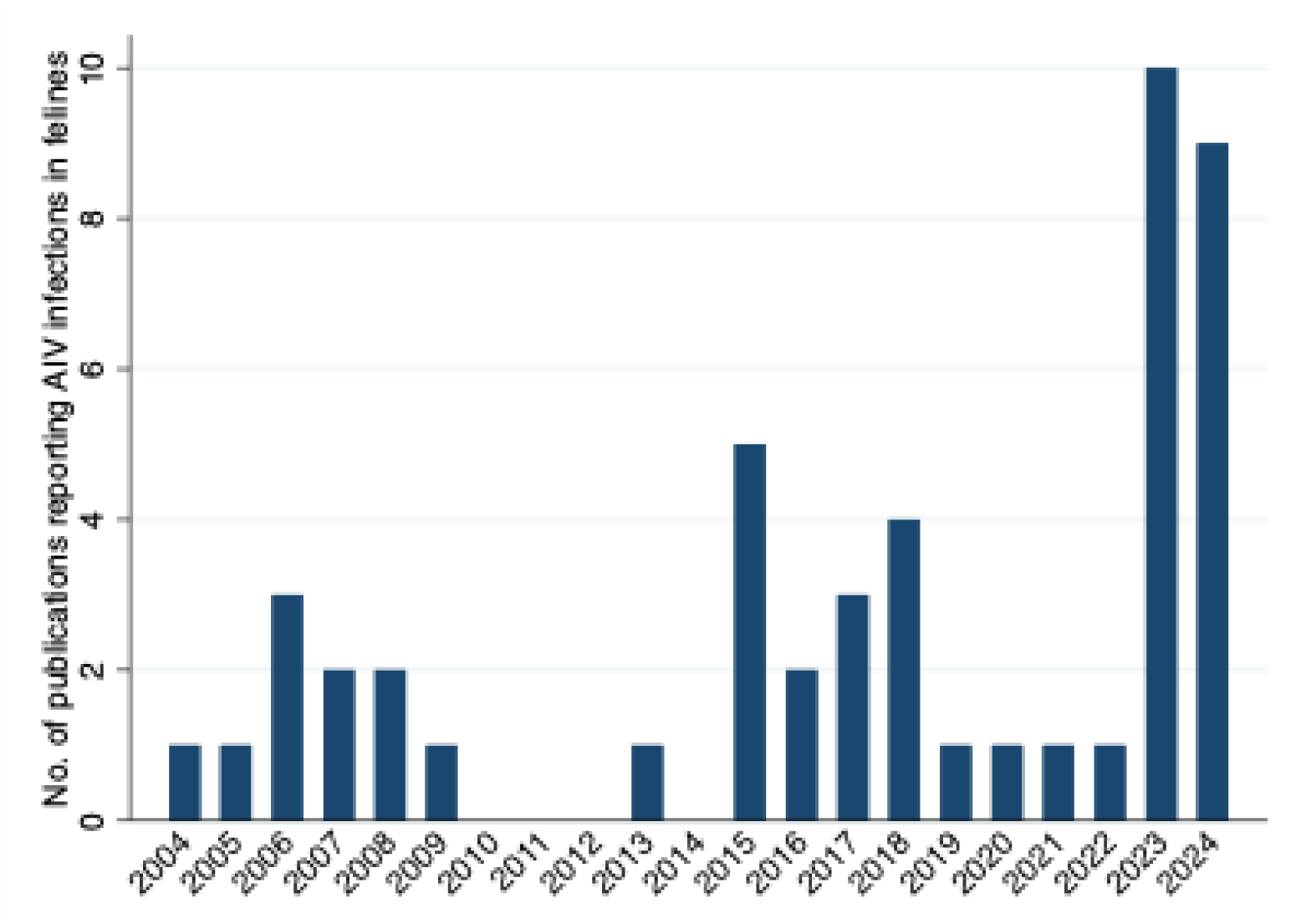
Number of peer-reviewed publications reporting avian influenza virus (AIV) infections in felines, each year from 2004 – 2024

**Figure 3.**
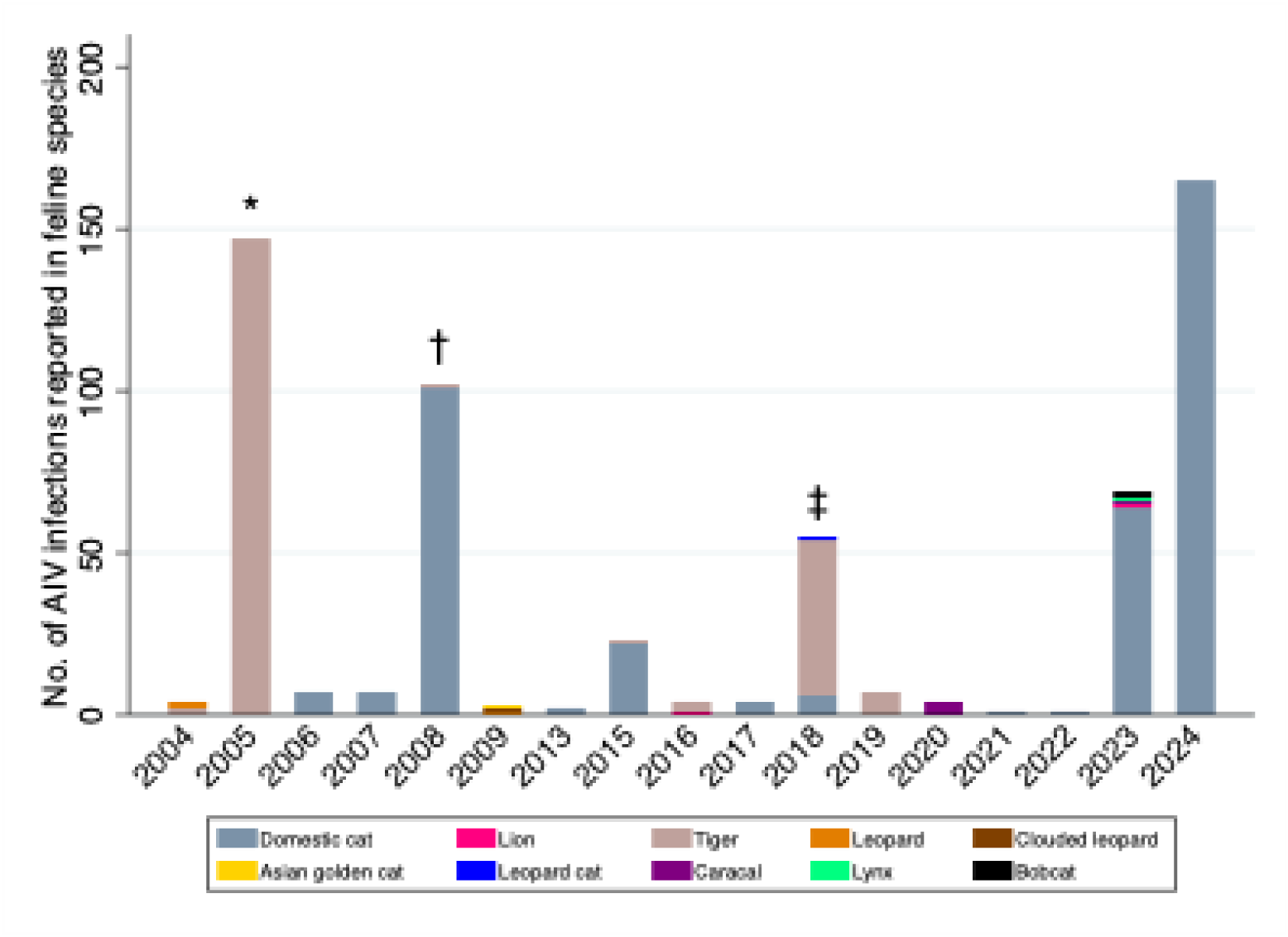
Number of avian influenza virus (AIV) infections in feline species reported each year in the peer-reviewed literature, 2004 – 2024. The year published may not be representative of the year in which each of the reported AIV infections occurred. *A total of 147 AIV-associated tiger deaths were reported in 2005 from an H5N1 outbreak in a Tiger Zoo in Thailand in 2004. †Approximately 100 of 500 domestic cats sampled in Indonesia were reported to be seropositive for H5N1 (the cited official reports of these tests are difficult to find). ‡Outbreaks of H7N2 among domestic cats occurred in multiple U.S. animal shelters but data published in the peer-reviewed literature do not include the number of infections and thus they are not included in this figure.

**Figure 4.**
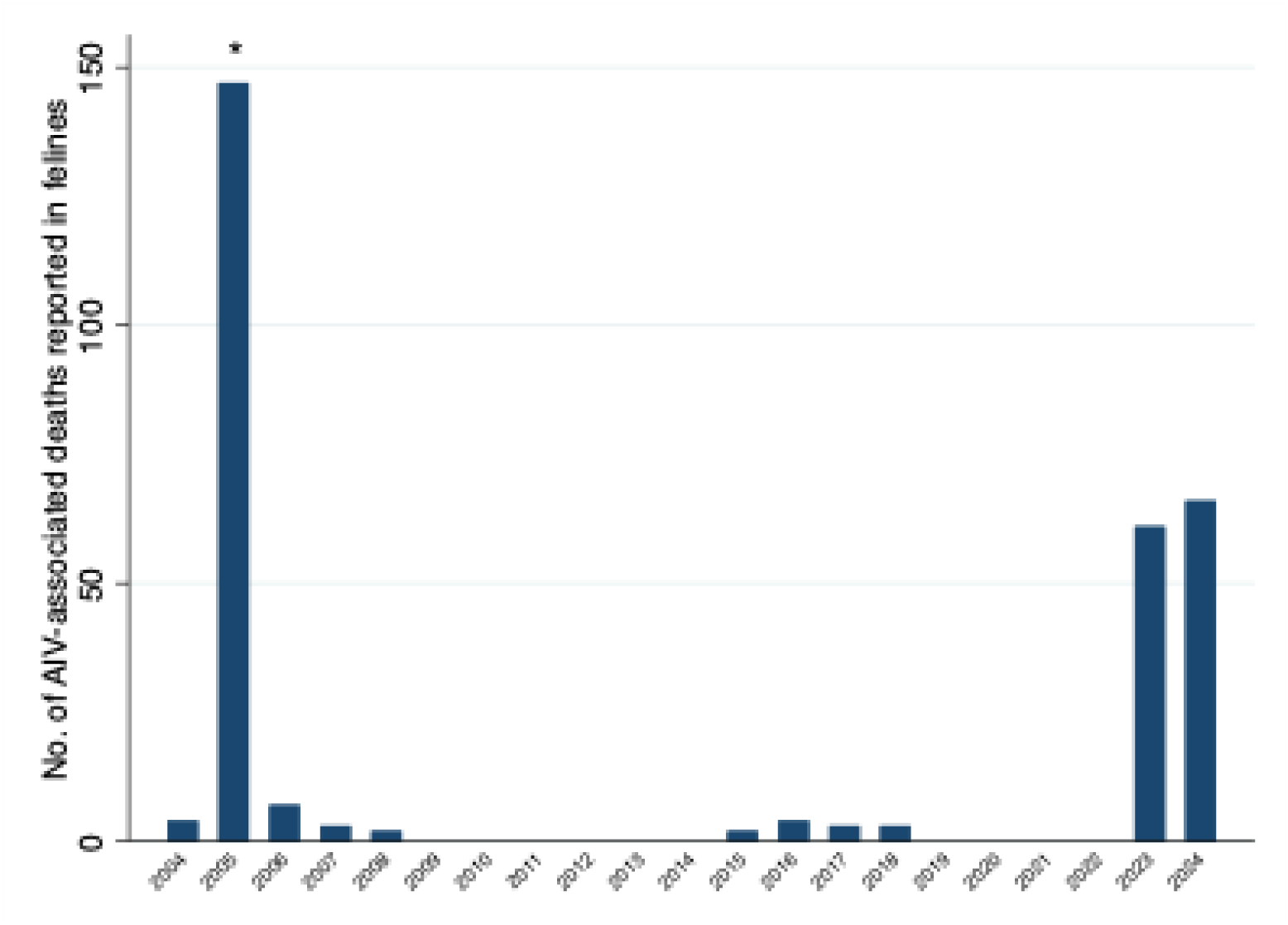
Number of avian influenza virus (AIV)-associated deaths in felines reported in the peer-reviewed literature, 2004 – 2024. The year published may not be representative of the year in which each of the reported AIV-associated deaths occurred. *147 AIV-associated tiger deaths were reported in 2005 from an H5N1 outbreak in a Tiger Zoo in Thailand in 2004.

### Avian influenza virus subtypes, clades, and fatality rates in felines

The overall case fatality rate among the reported RT-PCR-confirmed feline infections identified in our review was estimated to be 71.3%. Among the avian influenza virus infections reported in felines, 92.3% were identified as highly pathogenic avian influenza (HPAI) and 7.7% as low pathogenic avian influenza (LPAI). Among the RT-PCR-positive feline infections, HPAI accounted for 99.7% of the deaths.

Among all RT-PCR-confirmed feline infections, 98% (414/422) were specifically identified as HPAI H5N1, which had an overall case fatality rate of 71%. Among the H5N1 cases, 33.8% (140/414) were identified as clade 2.3.4.4b, of which 96.4% (135/140) were domestic cats. Among domestic cats, the overall PCR-confirmed case fatality rate was 52.8% for H5N1, and 89.6% for H5N1 clade 2.3.4.4b. See Table 2 for the remaining avian influenza virus subtypes and clades, including their respective case fatality rates, reported in felines.

**Table 2.** RT-PCR confirmed case fatality rate of avian influenza virus subtypes and clades identified in felines from 2004 – 2024.

| HPAI subtype | Case fatality rate |  | Clade | Case fatality rate |  |
| --- | --- | --- | --- | --- | --- |
|  | Feline | Domestic cat |  | Feline | Domestic cat |
| H5N1 | 71% | 52.8% | Clade 1 | 0% | 0% |
|  |  |  | Cade 2.2 | 100% | No infections |
|  |  |  | Clade 2.3.2 | 0% | No PCR+ cases |
|  |  |  | Clade 2.3.2.1c | 100% | No infections |
|  |  |  | Clade 2.3.2.1b | 100% | No infections |
|  |  |  | Clade 2.3.2.1e | 100% | No infections |
|  |  |  | Clade 2.3.4.4b | 90% | 89.6% |
|  |  |  | Clade 2.3.4.4e | 100% | No infections |
| H3N8 | 0% | 0% |  |  |  |
| H5N6 | 100% | 100% |  |  |  |
| H7N2 | 50% | 50% |  |  |  |
| H9N2 | 100% | 100% | C1.2 | 100% | 100% |

Influenza A H5N1 clade 2.3.4.4b was first detected in felines in 2022, including a wild lynx during an outbreak among pheasants in Finland [28], and a domestic cat living near a duck farm in France [29]. All studies reporting feline infections occurring since that time have identified H5N1 clade 2.3.4.4b as the causative agent [1,2,4,30–39], aside from two serology studies that did not report the specific clade of the H5 virus identified [40,41].

Among the publications that described the symptoms and pathology of the reported feline infections, respiratory and neurological illness were the most common and often resulted in death. Blindness and chorioretinitis were also recently observed in two AIV-infected domestic cats exposed to the virus through drinking raw colostrum and milk containing high viral loads from dairy cattle infected with H5N1 clade 2.3.4.4b [2]. This clinical observation suggests that exposure route and dose of AIV might impact disease presentation and severity. Although avian influenza in felines is often severe and fatal [6,29], subclinical infections have been reported [3,4].

### Geographic distribution of avian influenza virus infections in felines

Infections among 10 felid species across 7 geographical regions of the world were reported (Figure 5). The majority of reports (50%) were from Asia (including Southeast Asia), followed by Europe (25%) and North America (16.7%) (Supplementary Figure 1).

**Figure 5.**
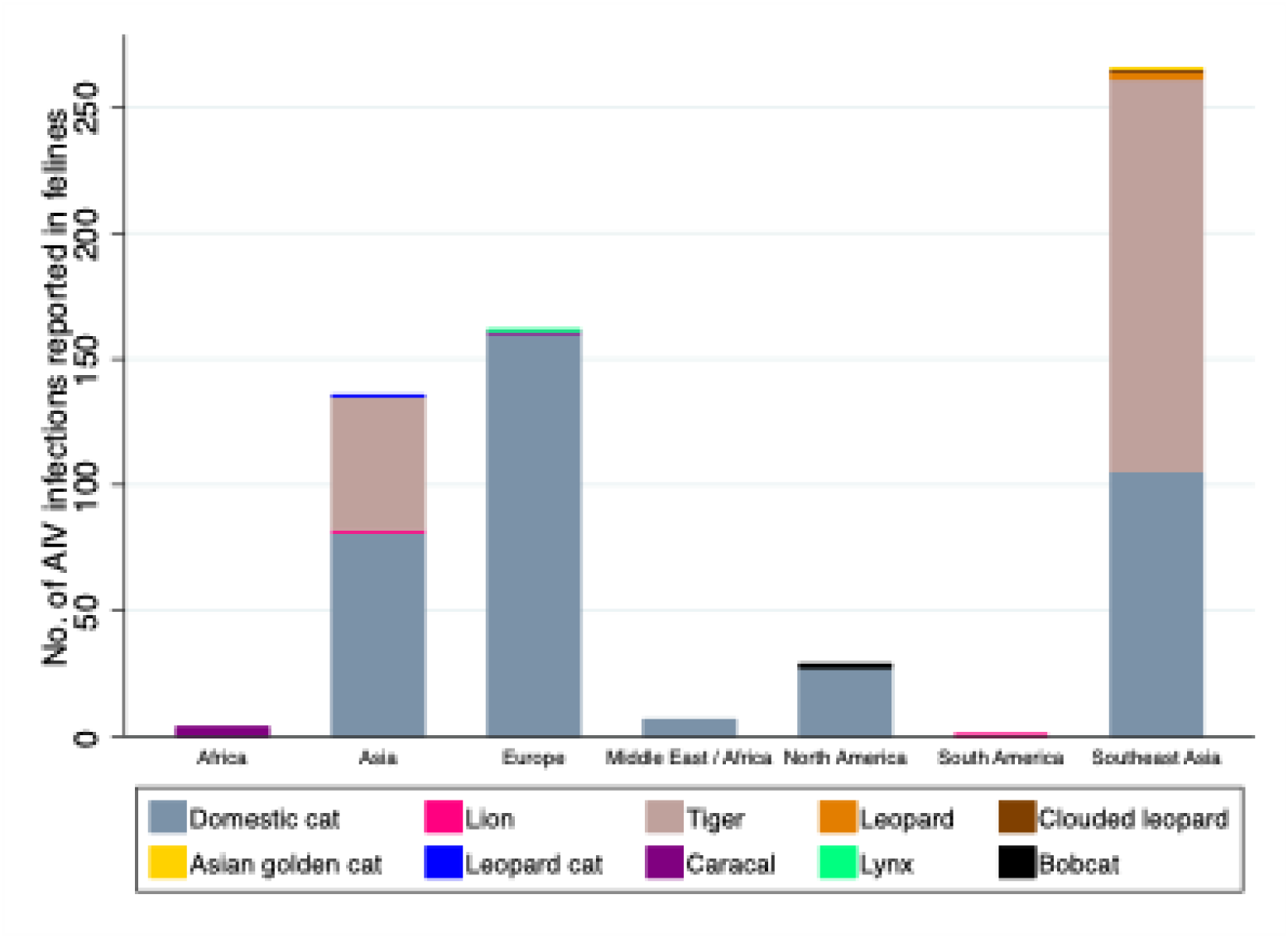
Geographical distribution of avian influenza virus (AIV) infections reported in feline species, 2004 – 2024

Feline infections of H5N1 clade 2.3.4.4b were reported in Finland, France, Poland, USA, Italy, Peru, and South Korea [1,2,4,28–37], including 5 species (135 domestic cats, two bobcats, and one lynx, caracal, and lion). Zoos, animal shelters, and rural settings such as farms and private land were the most common settings where AIV infections in felines were reported to occur.

### Cross- and intra-species transmission of avian influenza virus in felines

Most of the reported AIV infections in felines were confirmed or suspected to be a result of bird-to-feline transmission. Eating dead pigeons, chickens, and other birds, as well as contaminated raw chicken feed, was often implicated [6,39]. Domestic farm cats fed raw colostrum and milk from AIV-infected dairy cattle in Texas have also been infected [2], demonstrating a new route of cross-species, mammal-to-mammal transmission. Two studies, not including experimental infections, reported probable intra-species (feline-to-feline) transmission of AIV, including tiger-to-tiger [7] and domestic cat-to-cat [8] transmission.

Feline-to-human transmission of AIV has also been documented [7–9]. The first study to report feline-to-human transmission involved an outbreak of H5N1 among captive tigers in Thailand, after which two humans showed evidence of seroconversion [7]. The second involved a large outbreak of H7N2 among domestic cats in an animal shelter in New York, where two humans were infected, including a visiting veterinarian who collected samples from infected cats [8], and an animal shelter worker exposed to the infected cats [9]. Culture-positive air and surface samples were collected from the cat quarantine facility, demonstrating potential airborne and indirect contact (fomite-mediated) transmission [42]. An experimental animal study [6] of this particular H7N2 strain reported that ∼500 cats were infected in the animal shelters in New York, but we could not identify the primary source of this information.

## Discussion and Conclusion

Through our systematic review, we identified 607 avian influenza virus infections in felines, including 302 associated feline deaths, reported in the English scientific literature from 2004 – 2024. The reports represent cases from 7 geographical regions, including 18 countries and 12 felid species. The observed increase of AIV infections in felines over the past several years is commensurate with the rapid emergence of H5N1 clade 2.3.4.4b in other mammalian species. H5N1 clade 2.3.4.4b represents a variant in the hemagglutinin serotype 5 gene of IAV which became the dominant IAV H5 serotype among poultry in 2020 [43]. Clade 2.3.4.4b is the causative agent responsible for the observed spike in AIV infections among domestic cats in 2023 and 2024, involving at least 7 countries.

To date, HPAI H5N1 has been identified in at least 26 wild mammalian species in the U.S. [44]. As with many zoonotic pathogens, morbidity and mortality from AIV differs by species and virus subtype. In felines, HPAI results in an acute and often fatal course of illness characterized by atypical pneumonia, fever, encephalitis, and multisystem organ failure [2,31]. H5N1 clade 2.3.4.4b was first reported in felines in 2022, and among the feline infections reported in the literature thus far, it has yielded an extremely high case fatality rate of 90%. We suggest that this an overestimate due to the lack of testing and serosurveys among domestic cats, which accounted for 96.4% of the reported feline cases of clade 2.3.4.4b.

The most prominent symptom of HPAI in felines is acute encephalitis, with neuro invasion often resulting in ataxia, blindness, paralysis, and eventual death. The clinical presentation of encephalitis due to avian influenza is dramatic enough that it is often confused for rabies, likely leading to an underreporting of cases. This pattern is additionally observed in other carnivorous species, such as racoons, foxes, and coyotes, with acute encephalitis being the most common clinical finding [33]. Most surviving animals will have long term sequelae, particularly resulting in ataxia and seizures. As observed in cats [2], the encephalitis in wild carnivores often results in widespread necrosis evident upon examination of gross pathology, with the presence of cerebral necrosis [33]. Canines are also susceptible to AIV infection, as demonstrated by sporadic cases and deaths [14,21,45,46], as well as a recent H5N1 seropositivity rate of 2% among Washington hunting dogs with intense exposure to waterfowl [47].

Of particular concern is the increase of mammal-to-mammal transmission of avian influenza, indicating further adaptation to mammals. Recently, outbreaks of avian influenza have been reported in a new mammalian host: dairy cattle [2,37,48,49]. This is unusual considering the typical mammalian species affected and the transmission routes previously observed. Much of the infections in mammals have been reported in carnivores or swine, which are omnivorous. The transmission to herbivores is interesting, as avian influenza is often foodborne in mammalian hosts, and tends to result from a new host eating an infected host. The infection of ruminants rules out the predation or scavenging route of transmission in this case and suggests that other routes of transmission are occurring, in addition to cattle-to-cattle transmission. Exactly how avian influenza spilled over into dairy cattle is unknown [37,50]. It is interesting that dairy cattle outbreaks have been uniformly reported as H5N1 clade 2.3.4.4b and have been confined to the U.S., which is not the case for other domestic species, including domestic cats. In our review, we identified avian influenza virus infections in felines from 18 countries, of which 8 countries involved H5N1 clade 2.3.4.4b in domestic cats. We speculate that this geographic difference can be partly explained by route of transmission, as cats are carnivorous and prey on species susceptible to avian influenza such as birds and rodents. For example, a serologic study covering a wide range of African wildlife in Namibia demonstrates an association between diet and prior infection with influenza A virus [25]. In total, 21% (15/72) of carnivore-derived serum samples, representing brown and spotted hyenas, honey badgers, black-backed jackals, bat-eared foxes, African wild Dogs, lions, leopards, cheetahs, and caracals, were seropositive for hemagglutinin. Likewise, 44% (11/25) of the species that commonly prey on birds, including jackals, caracals, and honey badgers, were seropositive [25]. In contrast, only 5% (2/39) of herbivorous species serum samples were seropositive for hemagglutinin [25]. These two cases consisted of a single wildebeest seropositive for both H4 and H11 and a single black rhinoceros weakly seropositive for H5 [25]. However, a more recent serologic study of other herbivorous species in Pakistan demonstrates high seroprevalence of HPAI H5, with 27% of goats and 34% of sheep testing seropositive for clade 2.3.2.1c, and 24% of goats and 31% of sheep testing seropositive for 2.3.4.4b [51]. This study also tested for H7 and H9, with 14% of goats and 37% of sheep seropositive for H7, and 17% of goats and 35% of sheep seropositive for H9.

This new finding sets the stage for further surveillance and characterization of avian influenza virus infections among these domestic species, especially since the first known HPAI cases in U.S. ruminant livestock were among newborn goats infected with H5N1 clade 2.3.4.4b in February 2024 [52] – shortly before outbreaks among U.S. dairy cattle were reported. The newborn goat cases were presumed to be the result of direct spillover from nearby infected poultry. However, specific routes of transmission (e.g., foodborne, airborne) should be explored.

Most of the reported AIV infections in felines were confirmed or suspected to be a result of eating dead pigeons, chickens, and other birds, as well as contaminated raw chicken feed. The first reported foodborne-related instances involved outbreaks of H5N1 in tiger and leopard zoos and breeding operations in Thailand [7,53], which resulted in a large number of tiger deaths in 2004 [7]. The outbreaks were suspected to have started via infected chickens served to the tigers. The tigers were primarily fed whole, raw chicken carcasses, and at least one chicken carcass tested positive for H5N1 during the investigations. A similar outbreak of H5N1 also occurred at a wildlife rescue center in Cambodia in 2004 [54]. In this outbreak, a multitude of animals became infected, most notably several feline species, as well as several species of carnivorous birds. All were reported to have been fed chickens, although it was not specified if the feeder chickens were provided dead or alive. Considering that the outbreak among the carnivores ceased when the chickens were withdrawn as food, it was deemed that the outbreak was likely foodborne. More recently, foodborne transmission was suspected in AIV outbreaks among domestic cats in Poland in summer 2023 [30,31]. Culture-positive avian influenza virus was derived from a chicken sample from the freezer of the owners who fed such meat to one of the infected cats. As the 2023 feline outbreak in Poland coincided with an in-country poultry outbreak of H5N1 clade 2.3.4.4b, initial suspicions may have centered on predation or interactions with live birds as a means of transmission. However, as the outbreaks involved both indoor and outdoor felines, foodborne transmission of AIV was considered. As a result, frozen poultry samples were tested and found to be positive by MDCK cell culture. Illumina sequencing was performed, and the frozen poultry samples were differentiated from feline isolates by a maximum of 2 nucleotides per strand [30]. Two additional foodborne-related outbreaks of H5N1 clade 2.3.4.4b occurred in South Korea cat shelters in summer 2023 [35,36]. Improperly sterilized duck meat was implicated in both outbreaks [36].

In addition to predation and consumption of contaminated meat, a new phenomenon has emerged where domestic cats are being infected with H5N1 through the consumption of contaminated raw milk from U.S. dairy farms. H5N1 detections have also been reported in house and deer mice in New Mexico, Colorado, and Washington [44], which could potentially serve as an additional route of transmission to domestic cats. Interestingly, cases of H5N1 clade 2.3.4.4b recently reported by the Colorado Health Department included two indoor-only domestic cats with no known exposure to infected animals [55]. This observation raises concerns regarding new and unknown transmission routes of AIV to domestic cats. The USDA reports ∼96 HPAI H5 detections in felines from 2022 to present [44], of which approximately 20 have been reported in the scientific literature. Thus, important clinical, demographic, and sequencing data are unknown or unavailable for the majority of U.S. feline cases.

Our review, and especially calculations such as case fatality rate, were limited by the underreporting of avian influenza infections in felines in the scientific literature. Due to underreporting, our case count is an underestimate of the actual impact of avian influenza in feline hosts. Felines are not typically monitored for avian influenza. When a domestic feline or captive wild feline arrives at a clinic presenting with symptoms, such as severe atypical pneumonia, fever, and neurological problems such as ataxia and blindness, testing for avian influenza is not regularly performed. As avian influenza is an unusual diagnosis, and symptomatic cats would typically be euthanized to contain spread or for humane reasons, testing is of little benefit to the individual patient. However, from a One Health perspective, testing is highly valuable and can provide insight into nearby avian influenza outbreaks, as well as the potential risk of zoonotic spillover.

Based on the avian influenza seropositivity of various felines in some of the studies we reviewed, we estimate that feline cases are markedly underreported in the literature. Additionally, testing feline patients for AIV is not a routine practice as treatment options are limited under most circumstances. While it is often required that cases of HPAI shall be reported to public health authorities, subtype and clade is often not confirmed. As a result, several publications do not report the AIV subtype or clade. However, recent and ongoing outbreaks of H5N1 clade 2.3.4.4b among dairy cattle in the U.S., representing a significant threat to farm cats [2], has prompted multiple states to begin post-mortem H5N1 testing among rabies-negative domestic cats. However, subclinical infections of H5N1 in domestic cats do occur [3], including asymptomatic infections of clade 2.3.4.4b [4]. Thus, we argue that wider surveillance among domestic cats is urgently needed to determine how widespread the virus is in order to appropriately assess the risk of spillover to humans and other animals. As feline-to-human transmission of AIV has been documented [7–9], and potential airborne and fomite-mediated transmission implicated [42], farm and free-roaming cat owners, veterinarians, zoo keepers, and animal shelter volunteers may have a heightened risk of AIV infection during epizootics among birds and mammals.

## Data Availability

All data produced in the present study are available upon reasonable request to the authors.

## Acknowledgments

This work was inspired by Dr. Kristen Coleman’s kitten, Tuna. We acknowledge the support of the Department of Global, Environmental, and Occupational Health, at the University of Maryland School of Public Health, and the Department of Veterinary Medicine at the University of Maryland, College of Agriculture and Natural Resources. We also acknowledge the support of the University of Maryland Baltimore, Institute for Clinical & Translational Research (ICTR) and the University of Maryland Strategic Partnership: *MPowering the State* (MPower).

