## Supplemental Material for "Avian Influenza Virus Infections in Felines: A Systematic Review of Two Decades of Literature"

**A Systematic Review of Two Decades of Literature**

Supplementary Table 1. Search strategy for our systematic review of avian influenza virus infections in felines published in the scientific literature from 2004-2024.

| **Database** | **String of Search Terms Used** | **Results** |
| --- | --- | --- |
| PubMed | “ (“Avian influenza”[tw] OR “avian flu”[tw] OR “bird flu”[tw] OR “HPAI”[tw] OR “LPAI”[tw] OR “H5N1”[tw] OR “A(H5)”[tw] OR “A(H5N1)”[tw] OR “A/H5N1”[tw] OR “H7N9”[tw] OR “A(H7N9)”[tw]) AND (“feline”[tw] OR “felines”[tw] OR “cat”[tw] OR “cats”[tw] OR “kitten”[tw] OR “kittens”[tw] OR “felid”[tw] OR “felids”[tw] OR “lion”[tw] OR “tiger”[tw] OR “jaguar”[tw] OR “leopard”[tw] OR “cheetah”[tw] OR “caracal”[tw] OR “wildcat”[tw] OR “jaguarundi”[tw] OR “ocelot”[tw] OR “oncilla”[tw] OR “tigrillo”[tw] OR “kodkod”[tw] OR “güiña”[tw] OR “margay”[tw] OR “southern tigrina”[tw] OR “serval”[tw] OR “lynx”[tw] OR “linx”[tw] OR “bobcat”[tw] OR “manul”[tw] OR “cougar”[tw] OR “puma”[tw] OR “catamount”[tw]) AND ("2004/01/01"[PDAT] : "2024/12/31"[PDAT]) “ | 207 |
| Scopus | TITLE-ABS-KEY ( ( "Avian influenza" OR "avian flu" OR "bird flu" OR "HPAI" OR "LPAI" OR "H5N1" OR "A/H5N1" OR "H7N9" AND "feline" OR "felines" OR "cat" OR "cats" OR "kitten" OR "kittens" OR "felid" OR "felids" OR "lion" OR "tiger" OR "jaguar" OR "leopard" OR "cheetah" OR "caracal" OR "wildcat" OR "jaguarundi" OR "ocelot" OR "oncilla" OR "tigrillo" OR "kodkod" OR "guina" OR "margay" OR "southern tigrina" OR "serval" OR "lynx" OR "linx" OR "bobcat" OR "manul" OR "cougar" OR "puma" OR "catamount" ) ) AND PUBYEAR > 2003 AND PUBYEAR < 2025 AND ( LIMIT-TO ( DOCTYPE , "ar" ) OR LIMIT-TO ( DOCTYPE , "le" ) ) AND ( LIMIT-TO ( LANGUAGE , "English" ) ) | 207 |

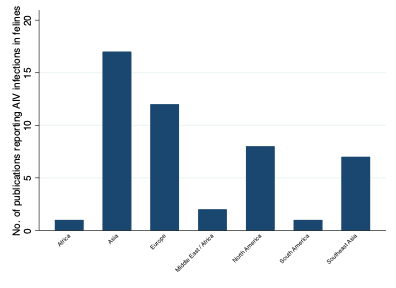

Supplementary Figure 1. Number of peer-reviewed publications reporting avian influenza virus (AIV) infections in felines, by geographic region of the world, 2004 – 2024
